# Health impacts of national and local air pollution control policies targeting electric generating units, mobile sources, and port activities in three US cities

**DOI:** 10.64898/2026.08.04.26359640

**Authors:** Haisu Zhang, Howard H. Chang, Ziqi Gao, Rohan R. D’Souza, Noah Scovronick, Philip K. Hopke, David Q. Rich, Armistead G. Russell, Stefanie Ebelt

## Abstract

**Objective:** Over the past decades, US policies intended to reduce air pollution emissions from electric generating units (EGUs), mobile sources (e.g., cars and trucks), and port activities have been implemented to improve air quality. This study aimed to estimate and compare counterfactual air pollution concentrations (i.e., concentrations that would have occurred without these policies) to observed concentrations, and then evaluate the health impacts of such policies in New York City, Los Angeles, and Atlanta from 2005 to 2019.

**Materials and Methods:** We obtained data on respiratory emergency department (ED) visits and cardiovascular disease ED visits that result in hospitalizations for the three cities from 2005-2019. Daily concentrations of fine particulate matter (PM_2.5_), criteria gases [carbon monoxide (CO), nitrogen dioxide (NO_2_), sulfur dioxide (SO_2_), and ozone (O_3_)], and 1-in-3-day measured concentrations of PM_2.5_ components and PM sources estimated using positive matrix factorization were acquired from six monitoring sites in the three cities. To estimate health impacts of selected EGU, mobile, and port policies we estimated: 1) counterfactual daily pollutant concentrations at each of the 6 city-sites; 2) associations between daily pollutant concentrations and rates of cardiorespiratory visits using city-site specific multi-pollutant Poisson models; and 3) the percent of cardiorespiratory visits prevented by the implementation of the selected policies, through applying observed and counterfactual concentrations to the fitted health models.

**Results:** Air quality policies were estimated to reduce ambient pollutant concentrations across the three cities, with median PM_2.5_ reductions of 27%-62% due to all policies combined during 2005-2019. Changes in criteria-pollutant concentrations associated with the selected policies were estimated to avert 7.1% (95% UI: 5.4%, 8.9%) of respiratory visits in New York City, 2.4% (95% UI: 1.4%, 3.4%) in Los Angeles, and 4.5% (95% UI: 0.8%, 8.2%) in Atlanta. In addition, 2.6% (95% UI: 0.9%, 4.2%) and 1.2% (95% UI: 0.3%, 2.1%) of cardiovascular visits were averted in New York City and Los Angeles, while the estimate in Atlanta did not indicate cardiovascular visits averted.

**Conclusion:** The selected EGU, mobile-source, and port policies evaluated during 2005-2019 were estimated to reduce ambient pollutant concentrations and avert respiratory visits in all three cities and cardiovascular visits in New York City and Los Angeles.

## Introduction

Air pollution represents one of the most significant environmental threats to public health worldwide, accounting for an estimated 6.7 million deaths globally in 2019 and ranking as the fourth leading risk factor for mortality (1, 2). It is estimated that 99% of the world’s population resides in places where ambient air quality failed to meet World Health Organization Air Quality Guideline levels as of 2019 (3). Cardiorespiratory conditions impose a particularly large global burden, as in 2019, chronic respiratory diseases were the third leading cause of death responsible for 4.0 million deaths globally, with chronic obstructive pulmonary disease (COPD) as the primary driver of respiratory mortality at 3.3 million deaths (4). It is also estimated that 19.8 million people died from cardiovascular diseases (CVD) in 2022, accounting for around 32% of global deaths (5).

Recent epidemiological studies continue to strengthen our understanding of the cardiorespiratory impacts of air pollution. A global systematic review and meta-analysis found a 2.29% increase in cardiorespiratory mortality per 10-µg/m³ increase in PM_2.5_, with associations for respiratory causes of death being stronger than for cardiovascular causes (6). A 2024 study found that among adults aged 65 years or older, chronic exposure to PM_2.5_ may increase the risk of hospitalization for cardiovascular conditions, particularly ischemic heart disease, cerebrovascular disease, heart failure, and arrhythmia (7). In a 15-year population-based cohort study of 5.1 million adults in Canada, long-term exposures to PM_2.5_, nitrogen dioxide (NO_2_), and ozone (O_3_) were associated with incident COPD, with hazard ratios ranging from 1.03 to 1.07 (8). Importantly, multiple large-scale epidemiological studies have associated air pollution exposure below regulatory standards with increased cardiovascular and respiratory morbidity and mortality over periods ranging from hours to years in diverse populations and geographic settings (9–12).

Over the past decades, US policies targeting electric generating units (EGU), mobile sources, and port activities have been implemented to reduce air pollution emissions and thus improve air quality. Air pollution accountability research is the process of evaluating the effectiveness of such policies and interventions in reducing emissions, air pollutant concentrations, and exposure, and improving health outcomes, which is crucial for informing future policy decisions and fostering public trust (13, 14). Several recent studies have evaluated the health benefits of Clean Air Act Amendment policies. For example, Meng and colleagues estimated reductions in emergency department (ED) visits and hospitalizations following implementation of California’s Emission Reduction Plan for Ports and Goods Movement program (15). Zigler and colleagues estimated the rate of all-cause Medicare mortality and respiratory hospitalization associated with PM_10_ nonattainment designations during 1990-1995 (16). Of greatest relevance to the current work, Russell and colleagues assessed the impact of EGU and mobile emission policies on the rates of cardiorespiratory ED visits and hospitalizations in metropolitan Atlanta during 1999-2013 (17). Most recently, Pitiranggon et al. estimated the source-specific health benefits of PM_2.5_ and NO_2_ reductions in New York City during 2005-2019, reporting substantial benefits from reductions in coal-fired power plant, residual-oil, and traffic emissions (18). While these accountability studies observed considerable reductions in pollutant concentrations attributed to policy implementation, the extent of public health improvements attributed to these policies varied, highlighting the need for further research.

In this study, we quantified the health benefits of EGU, mobile source, and port activity air quality policies in New York City, Los Angeles, and Atlanta, building on our foundational accountability work in Atlanta, GA (17, 19) and New York (20–23). We hypothesized that policy-driven changes in ambient concentrations of criteria pollutants [PM_2.5_, carbon monoxide (CO), NO_2_, sulfur dioxide (SO_2_), and O_3_] would lead to fewer cardiorespiratory ED visits. Further, we examined whether specific PM_2.5_ components and sources would similarly lead to these health benefits. Understanding the health benefits across multiple cities with diverse emission sources and policy contexts may provide critical evidence to guide resource allocation and prioritization of pollution control efforts nationwide.

## Methods

### Air Pollution and Meteorology Data

Daily air pollution data were obtained from six US EPA Chemical Speciation Network (CSN) monitoring sites from January 1, 2005 through December 31, 2019. The sites included three locations in New York City (Bronx [IS52], Manhattan [Division Street], Queens [Queens College 2]), two in Los Angeles (North Main Street, Rubidoux), and one in Atlanta (South DeKalb). These locations have been previously examined in air quality and health studies by our research group (20–24). Daily concentrations of criteria air pollutants (24-hr average PM_2.5_, 1-hr maximum CO, NO_2_, and SO_2_, and 8-hr average O_3_) were collected at all sites. Additionally, 1-in-3 day concentrations of PM_2.5_ components [sulfate (SO_4_^2-^), nitrate (NO_3_^-^), ammonium (NH_4_^+^), elemental carbon (EC), and organic carbon (OC)] and source-apportioned PM_2.5_ previously estimated using positive matrix factorization analysis [secondary sulfate (SS), secondary nitrate (SN), diesel vehicles (DIE), gasoline vehicles (GAS), road dust (RD), and residual oil (RO)] (25–28) were obtained. The Atlanta site, designated as a National Core site, also measured comprehensive gaseous species including online volatile organic compounds (VOCs). Where gaseous species monitoring was unavailable at CSN locations, data from nearby monitoring sites were incorporated. CSN analytical and quality assurance procedures are described elsewhere (29).

Hourly surface average temperature and dew point temperature data were obtained from the nearest airport weather station for all sites except Division St. for which data from the National Weather Service site in Central Park was employed (Table S1).

### Counterfactual Pollutant Concentration Estimation

Target policies affecting EGUs, on-road mobile sources, and ports were identified based on prior studies and expert consultations (Figure S1). Counterfactual pollutant concentrations were then estimated by linking policy-driven emissions changes to ambient concentrations using generalized additive models (GAMs). The policy groups evaluated varied by city: EGU and mobile-source policies were evaluated in New York City and Atlanta, whereas mobile-source and port policies were evaluated in Los Angeles. EGU policies were not evaluated in Los Angeles, and port policies were not evaluated in New York City or Atlanta, because emissions from these respective sources were low. Methods for doing so have been described previously (30, 31), and are summarized below. Briefly, for EGU policies, daily continuous emissions and monitoring data were tracked over time, with regulatory assessments used to identify when specific policies impacted emissions changes. For mobile source and port policies, emissions were modeled using EPA’s MOVES model for New York City and Atlanta, and California’s EMFAC model for Los Angeles. GAMs were then developed for each pollutant at each city-site to link actual emissions to daily concentrations, including meteorological factors (temperature, relative humidity, wind speed, dew point temperature, precipitation, and sea level pressure) modeled using cubic splines, and indicator variables for day-of-week, day-of-year, and holidays. Models included interaction terms between ammonia (NH_3_) and SO_2_/NO_x_ emissions, and between NO_x_, SO_2_, and VOC emissions with maximum temperature to capture photochemical processes. Model performance was evaluated using R², mean bias, root mean square error, and 10-fold cross-validation. Counterfactual concentrations were then calculated as 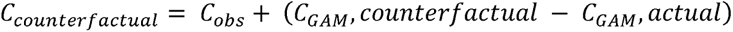, where *C_obs_* represents observed concentrations and *C_GAM_* terms represent GAM-predicted concentrations using counterfactual and actual emissions, respectively. Uncertainty was quantified through Monte Carlo simulations (n=500) using normal distributions for EGU emission ratios (based on base year mean and standard deviation) and ±50% uniform distributions for mobile and port emissions, with daily point estimates taken as simulation means and 95% uncertainty intervals from the 2.5^th^ and 97.5^th^ percentiles.

### Health Data

We obtained individual-level ED visit and hospitalization discharge data for 2005-2019 from New York State (Statewide Planning and Research Cooperative System), California (California Health and Human Services Agency), and Georgia (Georgia Hospital Association). We restricted analyses to patients who lived in ZIP codes located wholly or partially within 10 miles of a CSN monitoring site in Atlanta, New York City (Bronx, Manhattan, Queens) or Los Angeles (North Main Street, Rubidoux). For New York City, where 10-mile radius capture areas overlapped among the three monitoring sites, ZIP codes were assigned to the closest monitor based on centroid location. We defined respiratory disease and cardiovascular disease outcomes using selected International Classification of Diseases (ICD) discharge diagnosis codes (Table S2). During the study period, ICD codes transitioned from ICD-9 to ICD-10 in October 2015. Thus, we worked with clinical experts to ensure consistent outcome definitions across this transition. For the respiratory outcome, we included any ED visit directly discharged and ED visit resulting in hospitalization among all ages with a primary diagnosis for one of the selected ICD codes. For the cardiovascular outcome, we included only ED visits resulting in hospitalization among adults aged 19 years or older that had a primary diagnosis for one of the selected ICD codes.

### Statistical Analysis

#### Multi-Pollutant Health Models

To obtain an understanding of air pollution-health associations in the three cities over our study period, we first used multi-pollutant quasi-Poisson log-linear models to estimate overall associations between air pollutant concentrations and cardiorespiratory visits at each city-site:

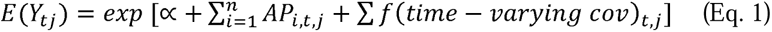

where *Y_tj_* represents daily visit counts for respiratory or cardiovascular outcomes on day t at site j, and *AP_i,t,j_* denotes air pollutant concentrations. We used 7-day (lags 0-6) moving average pollutant concentrations in models for respiratory visits and 4-day (lags 0-3) moving average pollutant concentrations in models for cardiovascular visits. Time-varying covariates included natural cubic splines with monthly knots for temporal trends, natural cubic splines (6 df) for temperature and dew point, and indicators for day-of-week and holidays. We selected these specifications based on sensitivity analyses that varied the lag periods, temporal-trend knot frequency, and degrees of freedom for temperature and dew point. These analyses were conducted using PM_2.5_-only models separately for cardiovascular and respiratory outcomes.

Four exposure sets were evaluated in separate models: 1) PM_2.5_ only; 2) five criteria pollutants (PM_2.5_, CO, NO_2_, SO_2_, O_3_); 3) four criteria gases plus five PM_2.5_ components (SO_4_^2-^, NO_3_ ^-^, NH_4_^+^, OC, EC); and 4) PM_2.5_ sources (SS, SN, GAS, DIE, RD, and RO). Exposure Set 2 was selected as the primary model because it provided broad coverage of criteria pollutants and daily exposure measurements. Pollutants were included only if available on ≥75% of days (daily measures) or ≥18% of days (1-in-3 day measures).

For each city and outcome, we estimated RRs corresponding to simultaneous one-IQR increases in all pollutants within the exposure set included in a given model. For New York City and Los Angeles, estimates were pooled across city-sites using inverse-variance weighting. IQR standardization facilitates comparison among pollutants measured on different concentration scales.

#### Health Benefits Assessment

We followed a two-stage framework to quantify the health impact of the selected air quality policies. The approach involved estimating pollution-health associations and applying them to both observed and counterfactual pollution concentrations to quantify policy-associated changes in cardiovascular or respiratory ED visits.

##### Stage 1: Health Effect Estimation

In Stage 1, we extended the multi-pollutant health models to include non-linear exposure terms:

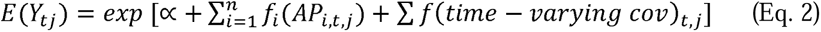

Where *f_i_*(*AP_i,t,j_*) denotes air pollutant concentrations modeled as natural cubic splines with three degrees of freedom.

##### Stage 2: Health impacts Quantification

In Stage 2, we calculated visits averted by comparing expected visit counts under counterfactual (policies not implemented) versus observed (policies implemented) scenarios. For each day, we estimated:

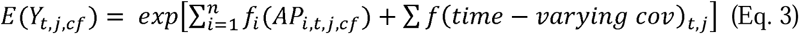

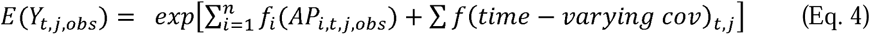

Daily visits averted were calculated as 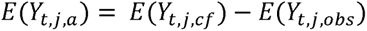. Positive values indicate visits averted under the observed scenario relative to the counterfactual scenario, whereas negative values indicate excess visits. We then calculated mean daily visits averted and percent of visits averted over specified time periods (full study period: 2005-2019; sub-periods: 2005-2007, 2008-2013, 2014-2016, 2017-2019).

We incorporated uncertainty through Monte Carlo simulations (n=5,000) accounting for: 1) uncertainty in daily counterfactual concentrations, and 2) uncertainty in model parameters. Daily counterfactual concentration standard deviations were estimated assuming point estimates and 95% bounds represented the 50th, 2.5th, and 97.5th percentiles of normal distributions. Regression coefficients were drawn from multivariate normal distributions defined by parameter estimates and variance-covariance matrices. Results are presented as medians with 95% uncertainty intervals (UIs) from simulation distributions.

Analyses were conducted separately for each city-site, outcome (respiratory, cardiovascular), exposure set (1–4), and applicable policy group (All, EGU, Mobile, Port). For New York City and Los Angeles, estimates were pooled across city-sites using inverse-variance weighting. All analyses were performed using R version 4.2.2.

## Results

### Changes in Air Pollutant Concentrations due to Policies

The observed and counterfactual ambient pollutant concentrations are shown in Table 1 and Tables S3 and S4. The selected air quality policies were estimated to reduce ambient pollutant concentrations across the three cities during 2005-2019, with some exceptions for O_3_. For example, median PM_2.5_ concentrations were reduced by 27%-62% due to all policies combined, with EGU policies driving larger PM_2.5_ concentration reductions in New York City (40%-44%) and Atlanta (29%) compared to mobile policies (6%-37% in New York City, 17%-20% in Los Angeles, 16% in Atlanta). In Los Angeles, mobile policies had slightly larger impacts than port policies (8%-15% reduction). Substantial reductions were also estimated for SO_2_ concentrations attributed to EGU policies (62%-77%) and port policies (58%-65%), and for PM_2.5_ from secondary sulfate (40%-90%), secondary nitrate (57%-79%), and gasoline vehicles (22%-71%) across city-sites.

**Table 1.**
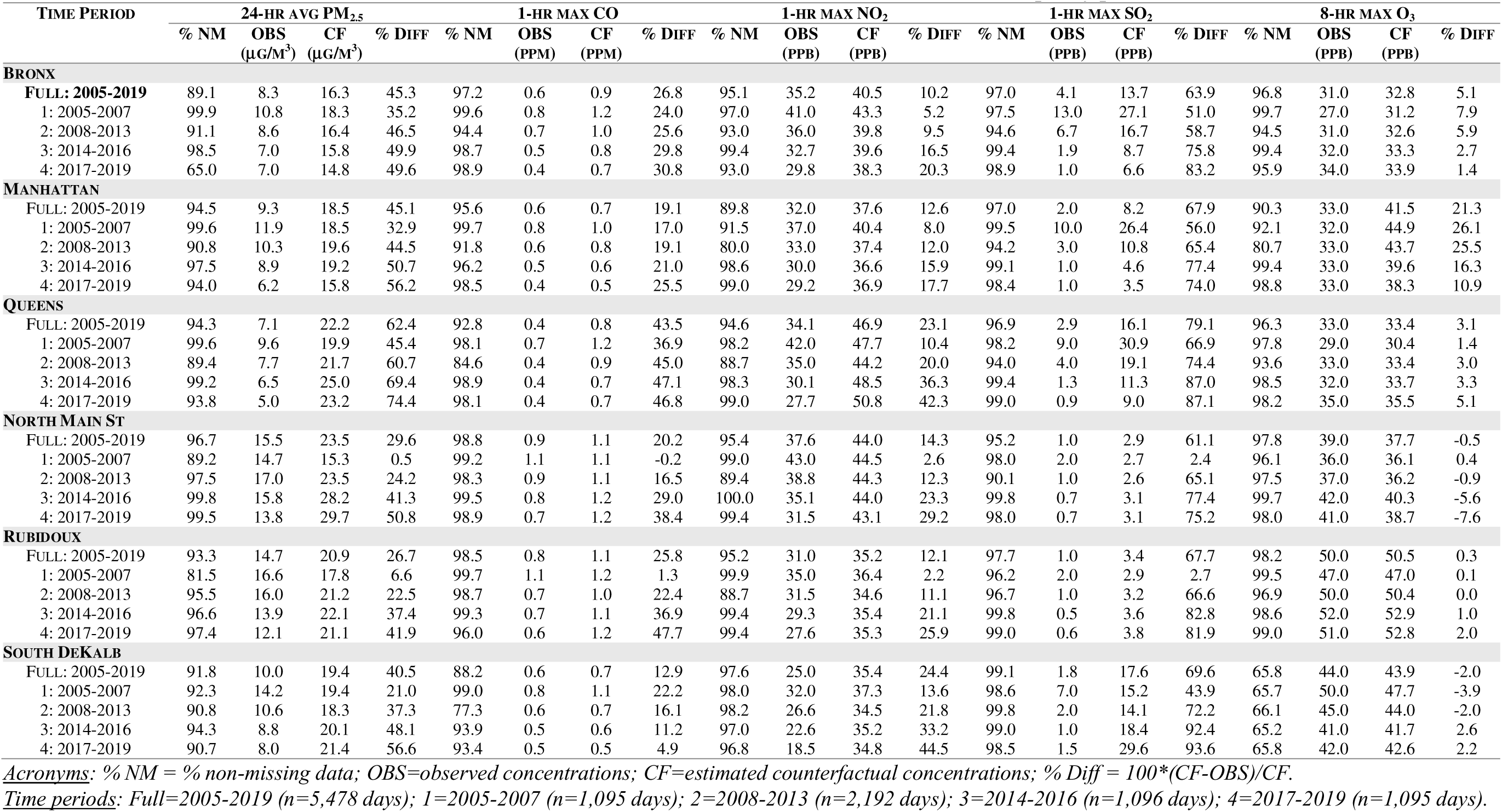
Criteria pollutants during full study period, 2005-2019, and over four sub-periods at six city-sites in New York City, Los Angeles, and Atlanta; comparing median observed and counterfactual concentrations estimated to have occurred in the absence of all selected air quality policies.

Policy impacts generally increased over time for most pollutants; for example, estimated PM_2.5_ reductions in Atlanta increased from 21% during 2005-2007 to 57% during 2017-2019, with similar temporal patterns observed in New York City (32%-45% to 50%-74%) and Los Angeles (<7% to 42%-51%). A notable exception was ozone, which increased in some locations and time periods. One possible explanation is the nonlinear response of ozone formation to NO_x_ emission reductions, particularly under cooler conditions with less sunlight.

### Cardiorespiratory ED Visits in Study Areas

Total and daily average numbers of cardiorespiratory ED visits during the 2005-2019 study period are shown in Table 2. There were 8,296,273 respiratory disease ED visits among patients of all ages across the six city-sites, with mean daily visit counts ranging from 116 visits/day (Rubidoux) to 433 visits/day (Bronx). For cardiovascular disease, there were 2,111,604 ED visits resulted in hospitalizations among patients aged 19 years and older, with mean daily visit counts ranging from 26 visits/day (Rubidoux) to 87 visits/day (Manhattan).

**Table 2.** Total and mean daily cardiorespiratory ED visits by corresponding city-site, 2005–2019.

| City | Monitoring site | Respiratory ED visits |  | Cardiovascular ED visits |  |
| --- | --- | --- | --- | --- | --- |
|  |  | Total number | Daily mean | Total number | Daily mean |
| New York City | Bronx | 2,374,290 | 433.4 | 450,913 | 82.3 |
|  | Manhattan | 1,704,695 | 311.2 | 475,430 | 86.8 |
|  | Queens | 1,276,766 | 233.1 | 395,921 | 72.3 |
| Los Angeles | North Main Street | 1,546,219 | 282.3 | 478,651 | 87.4 |
|  | Rubidoux | 636,977 | 116.3 | 143,093 | 26.1 |
| Atlanta | South DeKalb | 757,326 | 138.2 | 167,596 | 30.6 |
|  | **Overall** | 8,296,273 | 1,514.5 | 2,111,604 | 385.5 |

### Associations of Multi-Pollutant Exposures and Cardiorespiratory Visits

Associations between the four air-pollutant exposure sets and cardiorespiratory visits are presented in Table 3. In our primary model (Exposure Set 2), simultaneous one-IQR increases in all criteria pollutant concentrations were associated with higher rates of respiratory visits in all three cities: New York City (RR = 1.031, 95% CI: 1.018, 1.045), Los Angeles (RR = 1.023, 95% CI: 1.000, 1.047), and Atlanta (RR = 1.037, 95% CI: 1.013, 1.062). Similar levels of association were observed when considering PM_2.5_ mass alone (Exposure Set 1). The associations for criteria gases and PM_2.5_ components (Exposure Set 3) and PM_2.5_ sources (Exposure Set 4) were also positive in all three cities, but with wider confidence intervals, likely reflecting the fewer observations available because PM_2.5_ components and sources were measured every 3^rd^ day.

**Table 3.** Rate ratios (95% confidence intervals) for associations of multi-pollutant exposure sets and cardiorespiratory visits in three US cities, 2005-2019.

| City | Exposure Set 1: PM<sub>2.5</sub> Only | Exposure Set 2: Criteria | Exposure Set 3: Criteria | Exposure Set 4: PM<sub>2.5</sub> |
| --- | --- | --- | --- | --- |
|  |  | Pollutants | Gases + PM<sub>2.5</sub> Components | Sources |
| Respiratory Diseases |  |  |  |  |
| New York City | 1.027 (1.022, 1.033) | 1.031 (1.018, 1.045) | 1.011 (0.983, 1.040) | 1.010 (0.987, 1.033) |
| Los Angeles | 1.022 (1.019, 1.026) | 1.023 (1.000, 1.047) | 1.040 (0.993, 1.089) | 1.046 (1.020, 1.071) |
| Atlanta | 1.042 (1.029, 1.054) | 1.037 (1.013, 1.062) | 1.094 (0.998, 1.199) | 1.053 (0.996, 1.113) |
| Cardiovascular Diseases |  |  |  |  |
| New York City | 1.023 (1.018, 1.028) | 1.024 (1.013, 1.035) | 1.016 (0.991, 1.041) | 1.047 (1.024, 1.072) |
| Los Angeles | 1.006 (1.002, 1.010) | 1.025 (1.004, 1.047) | 1.032 (0.989, 1.078) | 1.024 (1.000, 1.049) |
| Atlanta | 1.010 (0.995, 1.025) | 1.011 (0.983, 1.041) | 0.944 (0.834, 1.068) | 0.987 (0.924, 1.055) |

All criteria air pollutants together (Exposure Set 2) were also associated with elevated cardiovascular visits in New York City (RR = 1.024, 95% CI: 1.013-1.035), Los Angeles (RR = 1.025, 95% CI: 1.004, 1.047). The Atlanta point estimate (RR = 1.011, 95% CI: 0.983, 1.041) was also positive, although its 95% confidence interval crossed the null. Associations for PM_2.5_ mass alone were similar in New York City and Atlanta but smaller in Los Angeles (RR = 1.006, 95% CI: 1.002, 1.010). For Exposure Sets 3 and 4, associations were positive but imprecise in New York City and Los Angeles. In Atlanta, however, the estimates were inverse for both Exposure Set 3 (RR = 0.944, 95% CI: 0.834, 1.068) and Exposure Set 4 (RR = 0.987, 95% CI: 0.924, 1.055).

### Respiratory Health Impacts of Air Quality Policies

The estimated numbers and percentages of respiratory visits averted due to air quality policies in all three cities during 2005-2019 are presented in Figure 1 and Table S5. For criteria pollutants (Exposure Set 2), we found 7.1% (95% UI: 5.4%, 8.9%), 2.4% (95% UI: 1.4%, 3.4%), and 4.5% (95% UI: 0.8%, 8.2%) of respiratory visits were averted in New York City, Los Angeles, and Atlanta, respectively. PM_2.5_-only models (Exposure Set 1) yielded similar but slightly lower estimates compared to the all-criteria pollutants models. Estimates of respiratory visits averted within the criteria gases and PM_2.5_ components models (Exposure Set 3) were also lower than the criteria pollutants model, with results close to zero in Los Angeles (0.6%, 95% UI: -0.7%, 1.9%). Estimates from the PM_2.5_ sources (Exposure Set 4) models were smallest for New York City and Atlanta, and for Los Angeles similar to the criteria pollutants estimate.

**Figure 1.**
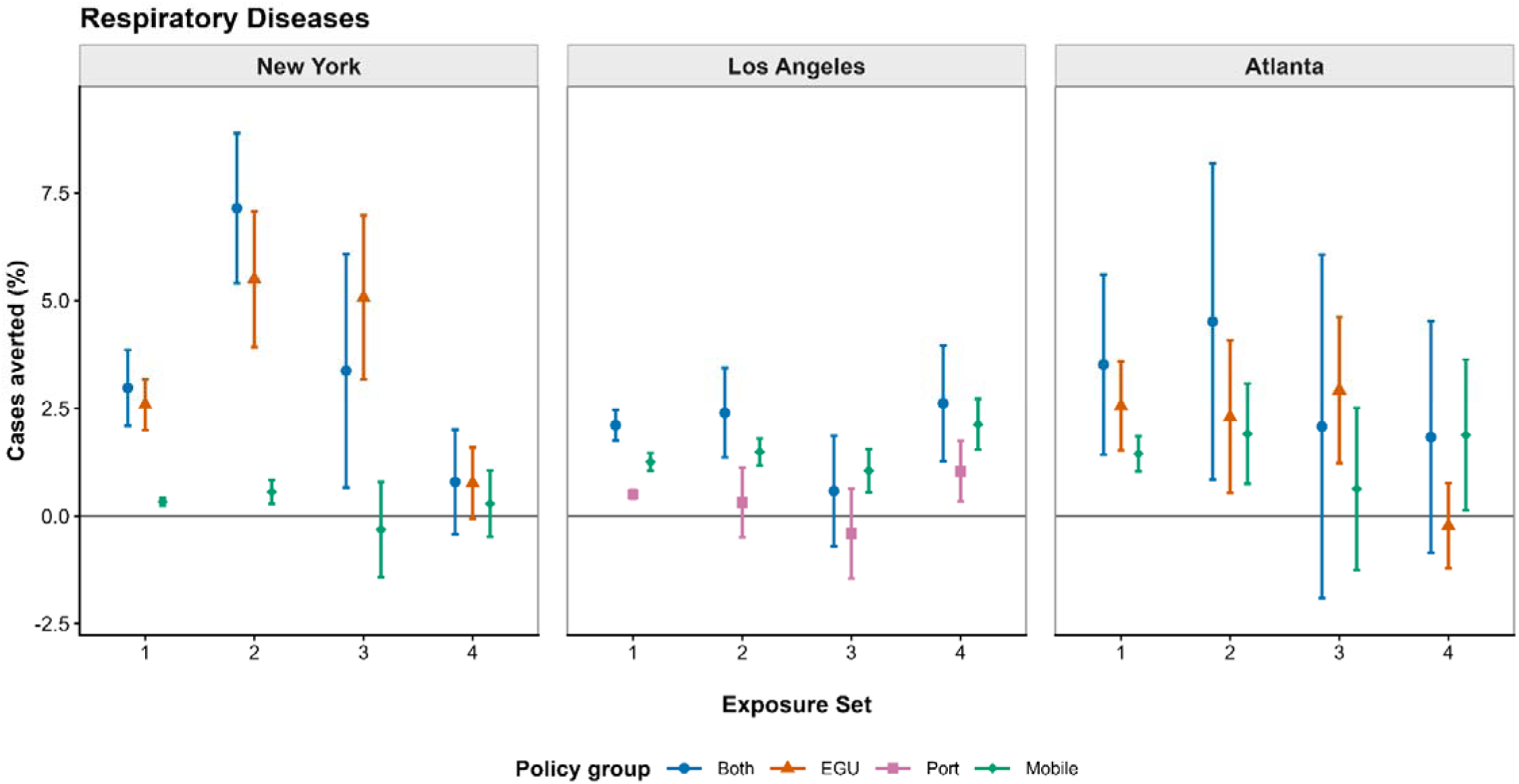
Estimated Percentage of Respiratory Disease Emergency Department Visits Averted by Policy Group, Exposure Set, and City, 2005–2019 Points indicate median estimates and whiskers indicate 95% uncertainty intervals derived from 5,000 Monte Carlo simulations. Exposure sets were: (1) PM_2.5_ only, (2) PM_2.5_ and gaseous pollutants, (3) PM_2.5_ components and gaseous pollutants, and (4) PM_2.5_ source factors. The city-specific results are pooled estimates of the percent of visits averted, which represent averages for populations living within 10-mile (roughly 314 square mile) areas represented by the city-sites. Positive values indicate visits averted; negative values indicate estimated increases in visits. EGU, electric generating unit; UI, uncertainty interval.

Comparing respiratory health impacts across policy groups, both EGU and mobile policies contributed to health benefits from criteria pollutant changes (Exposure Set 2), with EGU policies showing larger percentages of respiratory visits averted in New York City [5.5% (95% UI: 3.9%, 7.1%)] than mobile policies [0.6% (95% UI: 0.3%, 0.8%)], while Atlanta showed similarly sized estimates of respiratory visits averted for both policy groups. In Los Angeles, mobile policies were estimated to avert a larger percentage of respiratory visits [1.5% (95% UI: 1.2%, 1.8%)] than port policies [0.3% (95% UI: −0.5%, 1.1%)].

The estimated respiratory visits averted by policies increased over time in all three cities. For changes in criteria pollutant concentrations due to all policies, the percentages of respiratory visits averted in the most recent period (2017-2019) were 7.6% (95% UI: 5.8%, 9.4%) in New York City, 4.7% (95% UI: 3.1%, 6.2%) in Los Angeles, and 6.8% (95% UI: -0.6%, 14.1%) in Atlanta compared to 5.8% (95% UI: 3.5%, 8.2%) in New York City, -0.1% (95% UI: -0.2%, 0.1%) in Los Angeles, and 3.4% (95% UI: 1.2%, 5.6%) in Atlanta in the early period (2005-2007).

### Cardiovascular Health Impacts of Air Quality Policies

The estimated numbers and percentages of cardiovascular visits averted due to air quality policies in all three cities during 2005-2019 are presented in Figure 2 and Table S6. For criteria pollutants (Exposure Set 2), we found 2.6% (95% UI: 0.9%, 4.2%) and 1.2% (95% UI: 0.3%, 2.1%) of cardiovascular visits were averted in New York City and Los Angeles, respectively, and estimates were close to zero in Atlanta [-0.7% (95% UI: -5.5%, 4.1%)]. The results across the different exposure sets yielded largely similar results in all cities.

**Figure 2.**
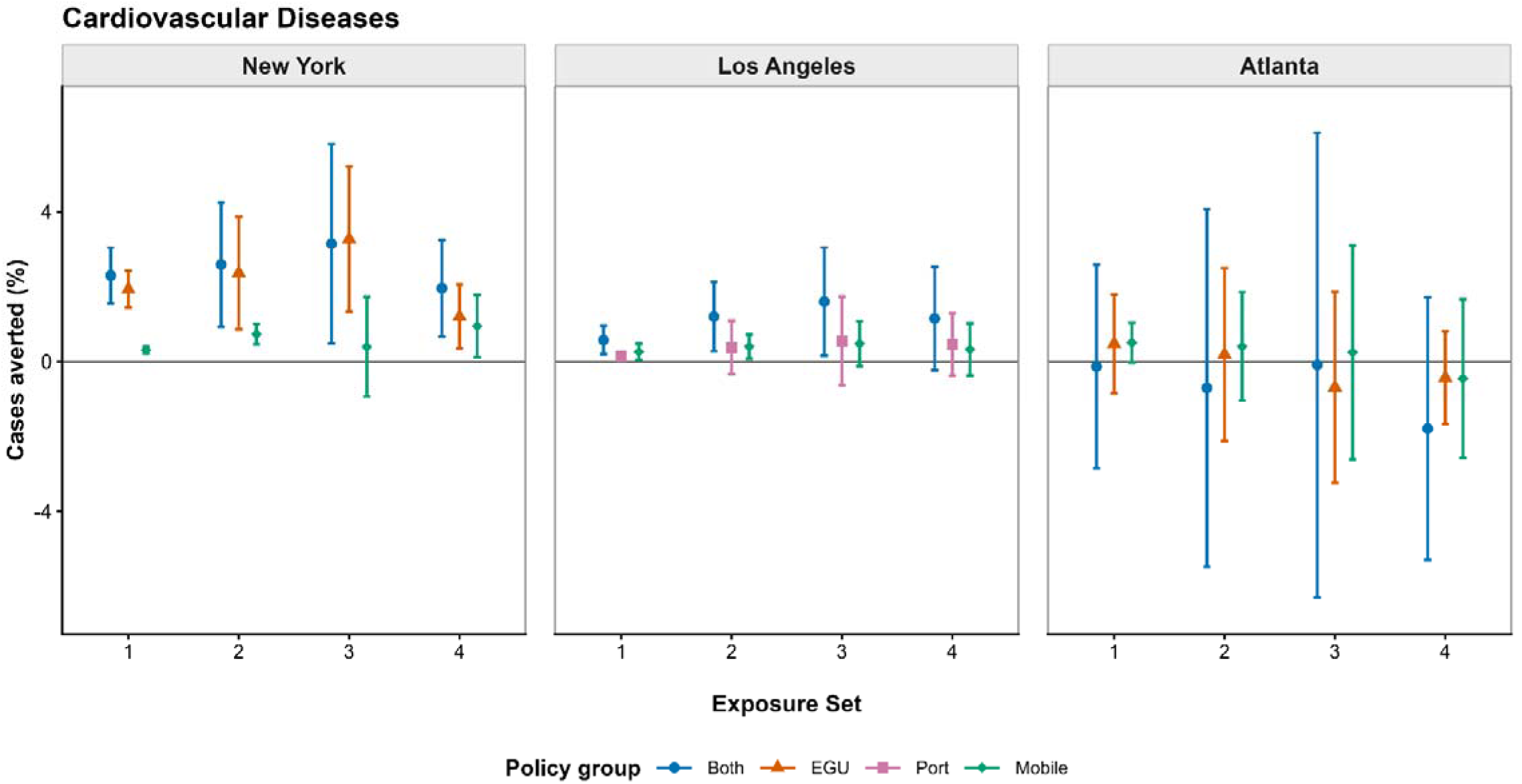
Estimated Percentage of Cardiovascular Disease Emergency Department Visits Averted by Policy Group, Exposure Set, and City, 2005–2019 Points indicate median estimates and whiskers indicate 95% uncertainty intervals derived from 5,000 Monte Carlo simulations. Exposure sets were: (1) PM_2.5_ only, (2) PM_2.5_ and gaseous pollutants, (3) PM_2.5_ components and gaseous pollutants, and (4) PM_2.5_ source factors. The city-specific results are pooled estimates of the percent of visits averted, which represent averages for populations living within 10-mile (roughly 314 square mile) areas represented by the city-sites. Positive values indicate visits averted; negative values indicate estimated increases in visits. EGU, electric generating unit; UI, uncertainty interval.

Both EGU and Mobile policies contributed to cardiovascular health benefits from criteria pollutant changes, with EGU policies showing larger impacts in New York City [2.4% (95% UI: 0.9%, 3.9%)] than mobile policies [0.7% (95% UI: 0.5%, 1.0%)]. In Los Angeles, mobile policies [0.4% (95% UI: 0.1%, 0.7%)] were estimated to have a similar impact on cardiovascular visits averted as port policies [0.4% (95% UI: -0.3%, 1.1%)].

Estimated percentages of cardiovascular visits averted by policies increased over time in New York City and Los Angeles and remained near 0% over all time periods in Atlanta. For criteria pollutant reductions from all policies, cardiovascular visits averted estimates were 3.6% (95% UI: 1.8%, 5.4%) in New York City and 2.1% (95% UI: 0.6%, 3.6%) in Los Angeles in the most recent period (2017-2019) compared to 1.6% (95% UI: -0.6%, 3.9%) in New York City and 0.2% (95% UI: 0.02%, 0.3%) in Los Angeles in the early period (2005-2007).

## Discussion

In this multicity accountability study, we found that air quality policies reduced ambient air pollutant concentrations across all three cities, with the estimated reductions increasing over time. We estimated that these air quality improvements averted respiratory emergency department visits and hospitalizations across all three cities, as well as cardiovascular hospitalizations in New York City and Los Angeles. Estimated health benefits were greatest during the later years of the study and for policies targeting electric generating units, demonstrating the growing public health benefits of emissions-control policies.

From the primary model (Exposure Set 2), we estimated that air quality policies averted 2.4%-7.1% respiratory visits across the three cities, with the largest percentage averted in New York City (7.1%), followed by Atlanta (4.5%) and Los Angeles (2.4%). In New York City, EGU policies accounted for the largest proportion of respiratory visits averted. This finding is consistent with Pitiranggon et al., who identified coal-fired power plant PM_2.5_ as the source with the largest concentration decline in New York City and estimated substantial associated reductions in asthma emergency department visits and hospitalizations among children aged 5-17 (18). The Atlanta results are similar to our previous accountability study using data from 1999-2013, which also estimated reduction of respiratory ED visits from policies (17, 19).

Estimated respiratory benefits varied across exposure sets. Compared with the PM_2.5_-only model (Exposure Set 1), the criteria-pollutant model (Exposure Set 2) estimated more than twice the benefit in New York City (7.1% for Exposure Set 2 vs. 3.0% for Exposure Set 1) and modestly greater benefits in Los Angeles (2.4% vs. 2.1%) and Atlanta (4.5% vs. 3.5%). This pattern suggests additional respiratory risks from gaseous pollutants beyond those captured by PM_2.5_ mass alone and is consistent with previous evidence linking short-term NO_2_ and SO_2_ exposure with respiratory morbidity and mortality (32–34). It also agrees with Pitiranggon et al., who estimated that a 40% reduction in NO_2_ in New York City avoided approximately 2,100 asthma emergency department visits (95% CI: 1,702-2,531) and 270 asthma hospitalizations (95% CI: 159-380) annually (18). The difference in estimated respiratory health benefits between Exposure Sets 2 and 1 was minimal in Los Angeles (2.4% vs. 2.1% of visits averted). One possible explanation is that baseline SO_2_ concentrations were relatively low in Los Angeles. For example, at the beginning of the study period, the SO_2_ concentration was 2.0 ppb at the Rubidoux site, compared with 13.0 ppb at the Bronx site in New York City.

Respiratory benefits estimated from Exposure Sets 3 and 4 generally had similar directions but wider uncertainty intervals compared to Exposure Sets 1 and 2, possibly because PM_2.5_ component and source measurements were available only every third day. It is notable that in New York City, including PM_2.5_ components or PM_2.5_ sources in the model reduced the health benefit sizes dramatically. A possible explanation is that the composition of PM_2.5_ mass changed over time, with earlier policies potentially reducing components less strongly associated with respiratory disease and the residual PM_2.5_ mixture then exhibited a different toxicity profile. Thus, using major PM_2.5_ components and sources data might deliver more accurate health impacts estimates on respiratory diseases compared to including PM_2.5_ mass.

For cardiovascular visits in the primary criteria-pollutant model (Exposure Set 2), air quality policies were estimated to avert cardiovascular visits in New York City and Los Angeles but not in Atlanta. This finding is supported by previous epidemiologic evidence linking short-term exposure to both particulate and gaseous criteria pollutants with cardiovascular outcomes. Studies have consistently reported associations between short-term criteria pollutant exposures and cardiovascular hospital admissions (35–39). For example, a nationwide study of adult Medicaid enrollees found that each 10-µg/m³ increase in the average PM_2.5_ concentration on the day of hospitalization and the preceding day was associated with a 0.9% increase in cardiovascular admission rates (95% CI: 0.6%-1.1%). A study in Beijing showed that 10 μg/m^3^ increase in daily mean NO_2_ concentration were associated with 1.89% increase in total cardiovascular disease mortality, with greater adverse effects on districts with larger population, and higher consumption of coal and more vehicles (38). In Atlanta, however, the estimated cardiovascular benefit was null. This result was generally consistent with our previous accountability study, which estimated that air quality policies prevented 2.3% of cardiovascular emergency department visits but reported a confidence interval that included the null (95% CI: −1.8% to 6.2%) (17). The wider uncertainty intervals in Atlanta may reflect its smaller study population and reliance on a single monitoring site, which may not fully represent population-level exposures across the metropolitan area.

The estimated cardiovascular benefits were greater for all criteria pollutants (Exposure Set 2) than for PM_2.5_ only (Exposure Set 1) in New York City (2.6% vs. 2.3% visits averted) and Los Angeles (1.2% vs. 0.6% visits averted). This modest increase in cardiovascular health benefits may reflect additional cardiovascular risks associated with gaseous criteria pollutants that are not captured by PM_2.5_ mass alone. The health benefit estimates for Exposure Sets 3 and 4 were less precise. Replacing PM_2.5_ mass with PM_2.5_ components (Exposure Set 2 vs. Exposure Set 3) or sources (Exposure Set 1 vs. Exposure Set 4) did not substantially alter the direction or magnitude of the estimated benefits. For example, in New York City, the estimated proportions of cardiovascular visits averted were similar between Exposure Sets 2 and 3 (2.6% vs. 3.1%) and between Exposure Sets 1 and 4 (2.3% vs. 2.0%). This suggests that PM_2.5_ mass may adequately represent the PM-related contribution to estimated cardiovascular benefits. Nevertheless, source-specific analyses remain valuable for identifying the emission sectors driving those benefits, as demonstrated by Pitiranggon et al. (18).

The estimated policy-related health benefits differed from the patterns observed in the multi-pollutant health models. For example, estimated cardiovascular benefits were generally smaller than respiratory benefits, which was not fully consistent with the RRs estimated in the multi-pollutant models. Similarly, although the multi-pollutant models showed comparable respiratory RRs for Exposure Sets 1 and 2, the estimated policy-related health benefits differed substantially between these exposure sets. Although the multi-pollutant models and policy health benefit models did not use identical specifications, these differences highlight the importance of counterfactual exposure estimates in evaluating the health benefits of air quality policies. In traditional multi-pollutant analysis, the joint RR represents a hypothetical simultaneous one-IQR increase in all included pollutants, a scenario unlikely to occur in practice. In contrast, our counterfactual approach modeled pollutant changes specific to each policy intervention, more closely reflecting the emission reductions actually achieved. This allowed for a more realistic evaluation of the health benefits associated with targeted air pollution control policies.

To better contextualize the public health impact of the selected air quality policies, we translated the estimated percent reductions into absolute numbers of healthcare encounters averted. Based on the average daily case count averted for Exposure Set 2 and the Both policy group, we estimated that policy-related reductions in criteria pollutant concentrations averted over 470,000 respiratory visits, and approximately 40,000 cardiovascular visits among adults in our three-city study populations from 2005-2019. These estimates suggest substantial public health benefits. Based on our data in which 86% of respiratory visits were ED visits not admitted to the hospital and 14% of respiratory visits were ED visits resulting in hospital admission. If these encounters were interpreted as visits avoided entirely, and applying published average costs for treat-and-release ED visits in large metropolitan areas in 2021 ($790) and hospital stays in 2012 ($10,400), the estimated respiratory visits averted could correspond to over $1 billion in healthcare costs avoided, with cardiovascular hospitalizations averted corresponding to an additional $400 million over the study period (40, 41). We note that these estimates are derived from city-site catchments (10-mile buffer around ground monitors) in three major metropolitan areas, do not represent complete citywide populations, and capture only two specific categories of health outcomes. Quantifying the broader health benefits across the United States would require a nationwide assessment.

Beyond the outcomes examined here, previous epidemiological studies have linked ambient air pollution to a wide range of additional adverse health outcomes. Prenatal exposure to criteria pollutants has been associated with adverse birth outcomes including preterm birth, low birthweight, and gestational hypertension (42, 43). Meanwhile, long-term PM_2.5_ exposure has been associated with accelerated cognitive decline and increased risk of dementia (44, 45). A meta-analysis also reported that a 10 μg/m³ increase in PM_2.5_ is associated with approximately a 16% increase in lung cancer risk (46). Accounting for these additional health outcomes, the corresponding public health and economic benefits would far exceed the $1 billion estimated in our study. Collectively, these findings suggest that air quality regulations function not only as environmental protection but as upstream preventive public health interventions. Benefit-cost analyses of the Clean Air Act Amendments have consistently demonstrated that the health benefits of pollution control programs substantially exceed their implementation costs, with net economic benefits estimated reaching trillions of dollars annually (47, 48). Overall, our findings suggest that regulatory investment in emission reductions represents a high-value strategy for reducing population-level disease burden.

There are several strengths of the present study. First, we extended the previous Atlanta analysis to a three-city assessment with diverse pollution sources, allowing comparisons of health benefits from policies in different settings. Second, the two-stage health benefits estimation framework across different exposure sets, along with the counterfactual pollution level estimates allowed more comprehensive estimation of health impacts from policies. The 15-year study period allowed us to evaluate cumulative policy impacts over an extended period. Third, we separately estimated the health effects to different groups of policies to provide policy-specific evidence that could inform targeted intervention for future regulations.

Several limitations should also be considered. The study populations were restricted to residents within 10 miles of CSN monitoring sites, which limited the generalizability of our findings. The counterfactual concentration estimation process relies on assumptions that cannot be fully validated. Statistical power was limited in certain model formulations, particularly when using 1-in-3 day PM_2.5_ component and source data or when dividing the study period into sub-periods for time-varying analyses, resulting in less stable estimates with wider confidence intervals. Finally, we used a single health model for the whole study period, which did not account for the potential changes in health associations between PM_2.5_ and cardiorespiratory outcomes over time. For example, prior work suggests that the toxicity of PM_2.5_ per unit mass may have increased for some health outcomes as its composition changed, with the long-term declines in relatively low-toxicity secondary inorganic species and increasing relative contributions from light-duty gasoline vehicles, diesel vehicle emissions and road dust in the remaining PM_2.5_ mass (26, 49–51).

## Conclusion

We estimated that EGU, mobile-source, and port policies reduced ambient concentrations of most pollutants and averted respiratory visits in all three cities and cardiovascular visits in New York City and Los Angeles. The use of a multi-pollutant accountability framework allowed us to estimate the health impacts of air quality policies more comprehensively. Our findings support sustained investment in emissions-control policies targeting EGU, mobile-source, and port sources to reduce cardiorespiratory morbidity.

## Supporting information

Supplementary Tables and Plots

## Data Availability

The health data analyzed in this study cannot be made publicly available because they contain sensitive information and are governed by privacy protections and data-use agreements. The authors are not authorized to redistribute these data. Aggregated results supporting the study findings are provided in the manuscript and supplementary materials.

## Acknowledgement

We are grateful for the support of this project by the Health Effects Institute (HEI) under award number 4986-RFA20-1A/21-9. In addition, health data utilized in this project were supported by grants to Emory University from the National Institute of Environmental Health Sciences (NIEHS) of the National Institutes of Health (NIH) under award numbers R01ES027892 and R01ES034175.

Source apportionment analyses in New York City were conducted as part of and with co-funding from Contract #’s 156226 and 125993 of the New York State Energy Research and Development Authority (NYSERDA) awarded to the University of Rochester. The content of this manuscript is solely the responsibility of the authors and does not necessarily represent the official views of HEI, NIH or NYSERDA.

We are also grateful for the support of the health data sources and their contributing hospitals that made this study possible. In particular, data on emergency department visits and hospitalizations were collected from patient-level hospital billing records acquired for: New York [New York State Department of Health, Statewide Planning and Research Cooperative System (SPARCS)]; California (California Health and Human Services Agency, Office of Statewide Planning and Development, now California Department of Health Care Access and Information); and Georgia (Georgia Hospital Association). The contents of this publication include data analysis, interpretation, conclusions derived, and the views expressed herein are solely those of the authors and do not represent the conclusions or official views of the data sources listed above. Authorization to release these data does not imply endorsement of this study or its findings by any of these data sources. The data sources, their employees, officers, and agents make no representation, warranty or guarantee as to the accuracy, completeness, currency, or suitability of the information provided here.

Finally, we thank Dr. Alvaro Alonso and Dr. Daniel Croft for providing clinical expertise and guidance on outcome definitions, including the selection of ICD codes for the health outcomes examined in this study.

