## Supplementary Tables and Plots for "Health impacts of national and local air pollution control policies targeting electric generating units, mobile sources, and port activities in three US cities"

**Table S1.** Meteorology monitoring sites paired with each CSN site, and data sources.

| City | Local CSN Site Name | Closest Paired Weather Station | Data Sources |
| --- | --- | --- | --- |
| New York | Bronx (IS52) | LaGuardia Airport (LGA) | NCEI |
| New York | Manhattan (Division St.) | National Weather Service site in Central Park | NCEI |
| New York | Queens College 2 | John F Kennedy International Airport (JFK) | NCEI |
| Los Angeles | North Main Street | Los Angeles International Airport (LAX) | CARB, National Solar Radiation Database |
| Los Angeles | Rubidoux | Ontario International Airport (ONT) | CARB, National Solar Radiation Database |
| Atlanta | South DeKalb | Atlanta Hartsfield International Airport (ATL) | NCEI |

*Data sources: NCEI = National Centers for Environmental Information (*[*https://www.ncei.noaa.gov/access/search/data-search/global-hourly*](https://www.ncei.noaa.gov/access/search/data-search/global-hourly)*); CARB =*

**Table S2.** Definitions of each health outcome of interest based on included ICD discharge diagnosis codes.

| Outcome | | ICD-9-CM Codes (for visits during 1/1/2005-9/30/2015) | ICD-10-CM Codes (for visits during 10/1/2015-12/31/2019) |
| --- | --- | --- | --- |
| Respiratory Diseases | |  |  |
|  | Upper Respiratory Infections | 460-465  (excludes code for acute bronchitis) | J00-J06 |
|  | Influenza | 487, 488  (includes broad set of codes for more sensitive definition) | J09, J10, J11  (includes broad set of codes for more sensitive definition) |
|  | Bacterial Pneumonia | 481, 482, 483.0, 483.1 | J13, J14, J15, J16, A48.1 |
|  | Culture-Negative Pneumonia | 485, 486 | J18 |
|  | Chronic Obstructive Pulmonary Disease | 491, 492, 496 | J41-J44 |
|  | Asthma | 493 | J45 |
| Cardiovascular Diseases | |  |  |
|  | Chronic Rheumatic Heart Disease | 393-396 | I05-I08 |
|  | Hypertension | 401-405, *not* 402.01, 402.11, 402.91, 404.01, 404.03, 404.11, 404.13, 404.91, 404.93  (excludes codes with heart failure) | I10-I13, I15-I16, *not* I11.0, I13.0, I13.2  (excludes codes with heart failure) |
|  | Ischemic Heart Disease | 410-414 | I20-I25 |
|  | Myocardial Infarction | 410 | I21, I22 |
|  | Cardiac Dysrhythmia | 427 | I46-I49  (includes code for cardiac arrest) |
|  | Heart Failure | 428, *and* 402.01, 402.11, 402.91, 404.01, 404.03, 404.11, 404.13, 404.91, 404.93  (includes codes for hypertensive heart disease with heart failure) | I50, *and* I11.0, I13.0, I13.2  (includes codes specific to HF, including hypertensive heart disease with heart failure) |
|  | Cerebrovascular Disease | 430-434, 436-438 | I60-I63, I65-I69 |
|  | Stroke | 430-432, 433.01, 433.11, 433.21, 433.31, 433.81, 433.91, 434.01, 434.11, 434.91, 436 | I60-I63 |

**Table S3.** PM_2.5_ components during full study period, 2005-2019, and over four sub-periods at six city-sites in New York City, Los Angeles, and Atlanta; comparing median observed and counterfactual concentrations estimated to have occurred in the absence of all selected AQ policies.

| Time Period | 24-hr avg SO_4_ | | | | 24-hr avg NO_3_ | | | | 24-hr avg NH_4_ | | | | 24-hr avg OC | | | | 24-hr avg EC | | | |
| --- | --- | --- | --- | --- | --- | --- | --- | --- | --- | --- | --- | --- | --- | --- | --- | --- | --- | --- | --- | --- |
|  | **%**  **NM** | **OBS**  **(μg/m^3^)** | **CF**  **(μg/m^3^)** | **%**  **Diff** | **%**  **NM** | **OBS**  **(μg/m^3^)** | **CF**  **(μg/m^3^)** | **%**  **Diff** | **%**  **NM** | **OBS**  **(μg/m^3^)** | **CF**  **(μg/m^3^)** | **%**  **Diff** | **%**  **NM** | **OBS**  **(μg/m^3^)** | **CF**  **(μg/m^3^)** | **%**  **Diff** | **%**  **NM** | **OBS**  **(μg/m^3^)** | **CF**  **(μg/m^3^)** | **%**  **Diff** |
| Bronx |  |  |  |  |  |  |  |  |  |  |  |  |  |  |  |  |  |  |  |  |
| Full: 2005-2019 | 19.5 | 1.48 | 2.91 | 34.3 | 19.5 | 0.83 | 1.12 | 18.8 | 19.5 | 0.67 | 1.70 | 36.6 | 19.4 | 1.65 | 1.79 | 4.5 | 19.4 | 1.00 | 2.81 | 57.9 |
| 1: 2005-2007 | 28.6 | 3.11 | 3.29 | 5.4 | 28.6 | 1.07 | 1.14 | 6.4 | 28.6 | 1.51 | 1.42 | -3.5 | 28.4 | 1.81 | 1.85 | 1.5 | 28.4 | 1.40 | 1.47 | 1.8 |
| 2: 2008-2013 | 9.2 | 2.28 | 3.04 | 21.0 | 9.2 | 0.72 | 0.90 | 14.4 | 9.2 | 1.14 | 1.51 | 20.6 | 9.2 | 1.32 | 1.37 | 3.5 | 9.2 | 1.29 | 1.79 | 27.5 |
| 3: 2014-2016 | 28.8 | 1.10 | 2.80 | 60.3 | 28.8 | 0.78 | 1.12 | 33.5 | 28.8 | 0.41 | 1.88 | 71.2 | 28.8 | 1.75 | 1.93 | 6.3 | 28.8 | 0.78 | 3.72 | 80.1 |
| 4: 2017-2019 | 21.6 | 0.76 | 2.66 | 67.5 | 21.6 | 0.77 | 1.27 | 38.3 | 21.6 | 0.25 | 1.87 | 83.3 | 21.2 | 1.60 | 1.81 | 9.2 | 21.2 | 0.69 | 6.17 | 86.9 |
| Manhattan |  |  |  |  |  |  |  |  |  |  |  |  |  |  |  |  |  |  |  |  |
| Full: 2005-2019 | 29.9 | 1.52 | 4.25 | 56.7 | 29.9 | 1.03 | 1.49 | 27.9 | 29.8 | 0.65 | 2.93 | 67.8 | 21.9 | 1.99 | 3.08 | 28.8 | 21.9 | 1.01 | 1.55 | 28.3 |
| 1: 2005-2007 | 28.8 | 2.94 | 4.09 | 23.3 | 28.8 | 1.65 | 1.83 | 8.1 | 28.7 | 1.56 | 3.10 | 41.9 | 26.8 | 2.34 | 2.88 | 17.5 | 26.8 | 1.16 | 1.18 | 0.7 |
| 2: 2008-2013 | 28.8 | 1.93 | 4.62 | 52.6 | 28.8 | 1.07 | 1.49 | 26.3 | 28.8 | 0.90 | 3.05 | 65.5 | 15.8 | 1.61 | 2.29 | 27.8 | 15.8 | 1.11 | 1.40 | 15.5 |
| 3: 2014-2016 | 31.5 | 1.13 | 4.24 | 63.6 | 31.5 | 0.86 | 1.43 | 34.9 | 31.0 | 0.44 | 2.96 | 80.8 | 25.5 | 1.78 | 3.12 | 39.6 | 25.5 | 0.95 | 1.70 | 42.7 |
| 4: 2017-2019 | 31.9 | 0.76 | 3.96 | 79.5 | 31.9 | 0.73 | 1.32 | 44.3 | 31.9 | 0.24 | 2.47 | 91.1 | 25.6 | 2.38 | 4.45 | 45.2 | 25.6 | 0.91 | 2.00 | 52.8 |
| Queens |  |  |  |  |  |  |  |  |  |  |  |  |  |  |  |  |  |  |  |  |
| Full: 2005-2019 | 31.4 | 1.43 | 3.58 | 50.7 | 31.3 | 0.85 | 1.40 | 33.3 | 31.1 | 0.58 | 2.57 | 68.6 | 24.9 | 1.41 | 2.48 | 26.0 | 24.9 | 0.67 | 1.31 | 41.1 |
| 1: 2005-2007 | 31.2 | 2.92 | 4.20 | 25.2 | 31.2 | 1.28 | 1.50 | 15.1 | 31.2 | 1.39 | 2.84 | 45.8 | 22.6 | 1.78 | 2.00 | 7.6 | 22.6 | 0.74 | 0.79 | 1.2 |
| 2: 2008-2013 | 30.6 | 1.80 | 3.91 | 47.3 | 30.6 | 0.88 | 1.37 | 31.7 | 30.6 | 0.77 | 2.68 | 65.4 | 22.7 | 1.39 | 1.84 | 20.7 | 22.7 | 0.74 | 1.11 | 27.0 |
| 3: 2014-2016 | 32.6 | 1.07 | 3.36 | 58.6 | 32.5 | 0.75 | 1.38 | 44.3 | 31.8 | 0.38 | 2.68 | 80.7 | 26.1 | 1.19 | 2.59 | 50.9 | 26.1 | 0.63 | 1.54 | 58.3 |
| 4: 2017-2019 | 31.9 | 0.73 | 2.98 | 70.5 | 31.8 | 0.64 | 1.39 | 49.4 | 31.3 | 0.21 | 2.26 | 87.7 | 30.1 | 1.43 | 4.14 | 65.0 | 30.1 | 0.55 | 1.96 | 71.0 |
| North Main St |  |  |  |  |  |  |  |  |  |  |  |  |  |  |  |  |  |  |  |  |
| Full: 2005-2019 | 24.8 | 1.37 | 3.37 | 46.4 | 24.7 | 2.46 | 4.59 | 39.2 | 24.4 | 0.75 | 2.28 | 51.3 | 24.4 | 2.14 | 3.68 | 38.9 | 24.4 | 0.64 | 0.82 | 20.9 |
| 1: 2005-2007 | 15.9 | 2.73 | 2.85 | 1.9 | 15.9 | 4.00 | 4.53 | 13.4 | 15.9 | 1.88 | 2.05 | 9.0 | 15.1 | 3.71 | 3.84 | 2.1 | 15.1 | 0.92 | 0.97 | 3.3 |
| 2: 2008-2013 | 23.1 | 1.54 | 3.45 | 47.8 | 23.0 | 2.82 | 4.93 | 37.1 | 23.0 | 0.94 | 2.35 | 49.8 | 23.1 | 1.97 | 3.08 | 35.8 | 23.1 | 0.68 | 0.88 | 22.0 |
| 3: 2014-2016 | 29.7 | 1.19 | 3.59 | 61.8 | 29.6 | 2.16 | 4.16 | 49.1 | 28.7 | 0.53 | 2.15 | 68.0 | 29.1 | 2.13 | 3.94 | 44.3 | 29.1 | 0.62 | 0.81 | 23.0 |
| 4: 2017-2019 | 32.1 | 1.09 | 3.25 | 70.0 | 32.1 | 2.14 | 4.72 | 51.5 | 31.3 | 0.54 | 2.31 | 69.0 | 31.5 | 2.22 | 4.68 | 50.6 | 31.5 | 0.47 | 0.65 | 26.1 |
| Rubidoux |  |  |  |  |  |  |  |  |  |  |  |  |  |  |  |  |  |  |  |  |
| Full: 2005-2019 | 31.7 | 1.29 | 2.17 | 27.7 | 31.7 | 3.37 | 5.57 | 23.8 | 31.5 | 1.11 | 2.08 | 25.7 | 30.8 | 0.75 | 1.00 | 18.4 | 30.8 | 2.68 | 3.56 | 16.5 |
| 1: 2005-2007 | 31.1 | 2.04 | 2.10 | 0.5 | 31.1 | 6.24 | 6.63 | 4.9 | 31.1 | 2.41 | 2.56 | 5.1 | 29.5 | 0.98 | 0.97 | -0.9 | 29.5 | 4.11 | 4.34 | 2.8 |
| 2: 2008-2013 | 32.3 | 1.48 | 2.28 | 28.7 | 32.3 | 3.85 | 5.56 | 25.3 | 32.3 | 1.40 | 2.06 | 28.7 | 32.2 | 0.85 | 0.98 | 11.3 | 32.2 | 2.51 | 3.51 | 20.1 |
| 3: 2014-2016 | 31.1 | 1.00 | 2.13 | 53.8 | 31.1 | 2.59 | 5.08 | 45.3 | 30.6 | 0.69 | 1.89 | 53.7 | 28.6 | 0.64 | 1.02 | 35.4 | 28.6 | 2.38 | 3.37 | 25.0 |
| 4: 2017-2019 | 31.7 | 0.96 | 2.05 | 54.8 | 31.7 | 2.57 | 5.33 | 50.6 | 31.4 | 0.67 | 1.86 | 63.5 | 31.6 | 0.55 | 1.01 | 45.4 | 31.6 | 2.58 | 3.36 | 19.7 |
| South DeKalb |  |  |  |  |  |  |  |  |  |  |  |  |  |  |  |  |  |  |  |  |
| Full: 2005-2019 | 27.9 | 1.62 | 4.63 | 49.3 | 27.9 | 0.33 | 0.58 | 40.7 | 28.6 | 0.45 | 1.63 | 57.2 | 27.9 | 2.32 | 6.99 | 55.7 | 27.9 | 0.61 | 0.85 | 19.2 |
| 1: 2005-2007 | 31.2 | 3.80 | 4.67 | 13.7 | 31.2 | 0.53 | 0.61 | 9.6 | 31.6 | 1.30 | 1.67 | 17.0 | 29.1 | 4.19 | 5.92 | 25.6 | 29.1 | 0.76 | 0.80 | 5.6 |
| 2: 2008-2013 | 25.6 | 2.02 | 4.42 | 49.5 | 25.6 | 0.33 | 0.55 | 35.4 | 27.5 | 0.58 | 1.53 | 54.5 | 25.6 | 2.13 | 5.65 | 53.7 | 25.6 | 0.68 | 0.82 | 16.4 |
| 3: 2014-2016 | 26.9 | 1.18 | 4.82 | 69.2 | 26.9 | 0.27 | 0.59 | 52.7 | 27.8 | 0.20 | 1.57 | 80.1 | 26.7 | 1.92 | 7.62 | 73.5 | 26.7 | 0.57 | 0.85 | 28.8 |
| 4: 2017-2019 | 30.0 | 0.83 | 4.80 | 82.7 | 30.0 | 0.21 | 0.60 | 62.9 | 28.8 | 0.16 | 2.03 | 91.4 | 32.3 | 1.93 | 11.83 | 83.4 | 32.3 | 0.46 | 0.91 | 49.1 |

*Acronyms: % NM = % non-missing data; OBS=observed concentrations; CF=estimated counterfactual concentrations; % Diff = 100*(CF-OBS)/CG.*

*Note: OBS concentrations were the same regardless of which policy group was assessed for CF estimation; for each city and pollutant, OBS statistics were paired with each CF estimate to facilitate comparison.*

*Note: Based on variable selection procedures in the counterfactual modeling in Chapter 4: 1) OBS and CF concentration estimates did not differ in some cases (e.g., for NY-Bronx, the model predicting SO_4_ concentrations only included SO_2_ emissions from EGU and other sources; for NY-Bronx and NY-Manhattan, models predicting NO_3_ concentrations only NOx emissions from mobile and other sources); and 2) OC and EC CF concentration estimates for all policies were the same as for mobile policies in some cases (OC and EC were not included in models of EGU policy impacts in NYC; EC was not included in models of port policy impacts in LA; and EC was not included in models of EGU policy impacts in ATL).*

**Table S4.** PM_2.5_ sources during full study period, 2005-2019, and over four sub-periods at six city-sites in New York City, Los Angeles, and Atlanta; comparing median observed and counterfactual concentrations estimated to have occurred in the absence of all selected AQ policies.

| Time Period | %  NM | 24-hr avg SS | | | 24-hr avg SN | | | 24-hr avg GAS | | | 24-hr avg DIE | | | 24-hr avg RD | | | 24-hr avg RO | | |
| --- | --- | --- | --- | --- | --- | --- | --- | --- | --- | --- | --- | --- | --- | --- | --- | --- | --- | --- | --- |
|  |  | **OBS**  **(μg/m^3^)** | **CF**  **(μg/m^3^)** | **%**  **Diff** | **OBS**  **(μg/m^3^)** | **CF**  **(μg/m^3^)** | **%**  **Diff** | **OBS**  **(μg/m^3^)** | **CF**  **(μg/m^3^)** | **%**  **Diff** | **OBS**  **(μg/m^3^)** | **CF**  **(μg/m^3^)** | **%**  **Diff** | **OBS**  **(μg/m^3^)** | **CF**  **(μg/m^3^)** | **%**  **Diff** | **OBS**  **(μg/m^3^)** | **CF**  **(μg/m^3^)** | **%**  **Diff** |
| Bronx |  |  |  |  |  |  |  |  |  |  |  |  |  |  |  |  |  |  |  |
| Full: 2005-2019 | 19.3 | 1.37 | 8.65 | 78.0 | 0.82 | 3.64 | 69.7 | 1.56 | 2.84 | 45.3 | 0.72 | 1.03 | 16.0 | 0.44 | 1.02 | 61.5 | 0.28 | 0.80 | 53.1 |
| 1: 2005-2007 | 28.2 | 4.33 | 9.05 | 52.4 | 1.05 | 3.22 | 50.4 | 1.18 | 2.16 | 45.1 | 0.97 | 1.04 | 6.7 | 0.42 | 0.45 | 2.5 | 0.72 | 1.21 | 29.2 |
| 2: 2008-2013 | 9.1 | 3.18 | 9.41 | 63.9 | 0.94 | 3.30 | 69.8 | 0.49 | 1.81 | 73.8 | 0.59 | 0.74 | 18.8 | 0.72 | 0.88 | 20.8 | 0.49 | 1.02 | 51.7 |
| 3: 2014-2016 | 28.5 | 0.45 | 8.46 | 91.1 | 0.58 | 3.87 | 69.3 | 2.10 | 3.65 | 41.6 | 0.64 | 1.02 | 39.3 | 0.41 | 1.13 | 65.2 | 0.15 | 0.60 | 70.3 |
| 4: 2017-2019 | 21.5 | 0.12 | 8.33 | 97.6 | 0.63 | 4.06 | 70.1 | 2.38 | 3.91 | 39.7 | 0.79 | 1.29 | 36.2 | 0.31 | 1.34 | 88.7 | 0.15 | 0.58 | 67.4 |
| Manhattan |  |  |  |  |  |  |  |  |  |  |  |  |  |  |  |  |  |  |  |
| Full: 2005-2019 | 21.9 | 1.51 | 8.79 | 79.2 | 0.45 | 9.12 | 90.1 | 0.75 | 2.28 | 65.5 | 0.89 | 2.01 | 44.8 | 0.24 | 0.41 | 33.0 | 0.49 | 0.49 | 0.0 |
| 1: 2005-2007 | 26.8 | 2.47 | 8.94 | 72.8 | 2.32 | 8.35 | 62.1 | 0.83 | 1.65 | 56.0 | 1.33 | 1.77 | 23.5 | 0.67 | 0.68 | 0.5 | 0.40 | 0.40 | 0.0 |
| 2: 2008-2013 | 15.7 | 2.62 | 9.85 | 73.1 | 0.49 | 8.05 | 88.0 | 0.28 | 1.52 | 78.8 | 0.85 | 1.51 | 40.7 | 0.19 | 0.27 | 21.2 | 0.78 | 0.78 | 0.0 |
| 3: 2014-2016 | 25.5 | 0.87 | 7.60 | 83.1 | 0.20 | 9.79 | 93.0 | 0.69 | 2.63 | 71.7 | 0.79 | 2.24 | 64.7 | 0.19 | 0.39 | 53.4 | 0.39 | 0.39 | 0.0 |
| 4: 2017-2019 | 25.6 | 0.68 | 8.63 | 91.4 | 0.11 | 10.32 | 95.2 | 1.48 | 3.69 | 63.9 | 0.81 | 2.95 | 71.0 | 0.23 | 0.51 | 56.7 | 0.38 | 0.38 | 0.0 |
| Queens |  |  |  |  |  |  |  |  |  |  |  |  |  |  |  |  |  |  |  |
| Full: 2005-2019 | 24.9 | 1.68 | 6.89 | 75.3 | 1.07 | 12.06 | 86.9 | 0.49 | 1.23 | 43.3 | 0.61 | 0.95 | 31.4 | 0.09 | 0.43 | 54.7 | 0.05 | 0.15 | 57.8 |
| 1: 2005-2007 | 22.6 | 3.51 | 6.74 | 46.4 | 1.26 | 9.37 | 83.6 | 0.62 | 1.12 | 38.8 | 1.07 | 1.24 | 11.4 | 0.54 | 0.55 | 1.0 | 0.11 | 0.14 | 25.9 |
| 2: 2008-2013 | 22.7 | 2.79 | 7.67 | 62.8 | 0.77 | 10.39 | 90.0 | 0.58 | 1.16 | 45.1 | 0.58 | 0.87 | 34.3 | 0.09 | 0.26 | 52.3 | 0.06 | 0.14 | 57.8 |
| 3: 2014-2016 | 26.1 | 0.70 | 6.46 | 85.4 | 0.87 | 13.36 | 90.1 | 0.37 | 1.37 | 71.6 | 0.55 | 0.92 | 40.9 | 0.08 | 0.42 | 85.0 | 0.04 | 0.15 | 72.4 |
| 4: 2017-2019 | 30.4 | 0.26 | 6.50 | 94.3 | 1.42 | 17.62 | 90.3 | 0.29 | 1.56 | 73.5 | 0.46 | 0.92 | 52.2 | 0.05 | 0.57 | 87.7 | 0.03 | 0.16 | 77.7 |
| North Main St |  |  |  |  |  |  |  |  |  |  |  |  |  |  |  |  |  |  |  |
| Full: 2005-2019 | 24.2 | 0.78 | 3.54 | 71.8 | 1.68 | 5.45 | 41.3 | 0.82 | 3.93 | 71.4 | 2.33 | 6.20 | 50.3 | 0.35 | 0.83 | 56.8 | NA | NA | NA |
| 1: 2005-2007 | 15.1 | 1.62 | 1.77 | 2.8 | 2.50 | 2.99 | 13.6 | 1.65 | 2.45 | 32.2 | 1.81 | 2.64 | 33.3 | 0.59 | 0.70 | 11.4 | NA | NA | NA |
| 2: 2008-2013 | 23.1 | 1.00 | 3.49 | 65.5 | 2.38 | 3.85 | 32.5 | 0.73 | 3.02 | 78.8 | 2.48 | 5.00 | 50.7 | 0.32 | 0.70 | 59.1 | NA | NA | NA |
| 3: 2014-2016 | 28.1 | 0.63 | 4.02 | 77.6 | 1.02 | 5.43 | 72.2 | 0.81 | 4.07 | 80.0 | 2.92 | 6.99 | 57.6 | 0.35 | 0.87 | 60.1 | NA | NA | NA |
| 4: 2017-2019 | 31.5 | 0.50 | 3.73 | 84.8 | 1.35 | 8.93 | 76.3 | 0.74 | 5.00 | 83.3 | 2.14 | 7.97 | 70.3 | 0.33 | 0.93 | 65.5 | NA | NA | NA |
| Rubidoux |  |  |  |  |  |  |  |  |  |  |  |  |  |  |  |  |  |  |  |
| Full: 2005-2019 | 29.8 | 0.73 | 2.74 | 65.9 | 2.94 | 7.94 | 40.1 | 1.09 | 1.67 | 22.4 | 2.21 | 2.75 | 15.9 | 0.56 | 0.60 | 2.1 | NA | NA | NA |
| 1: 2005-2007 | 28.9 | 1.38 | 1.42 | 2.8 | 7.08 | 7.38 | 6.8 | 0.76 | 0.94 | 13.8 | 2.10 | 2.34 | 8.5 | 0.46 | 0.46 | 1.0 | NA | NA | NA |
| 2: 2008-2013 | 31.8 | 1.00 | 2.88 | 61.0 | 3.50 | 6.74 | 38.1 | 1.16 | 1.47 | 21.8 | 1.79 | 2.25 | 19.5 | 0.51 | 0.53 | 2.8 | NA | NA | NA |
| 3: 2014-2016 | 26.3 | 0.49 | 3.20 | 81.3 | 1.67 | 8.41 | 71.0 | 1.25 | 1.86 | 35.2 | 2.44 | 2.97 | 20.0 | 0.69 | 0.72 | 2.5 | NA | NA | NA |
| 4: 2017-2019 | 30.2 | 0.27 | 2.87 | 89.0 | 1.52 | 10.97 | 74.3 | 1.25 | 2.25 | 44.1 | 3.09 | 3.89 | 22.4 | 0.72 | 0.73 | 2.8 | NA | NA | NA |
| South DeKalb |  |  |  |  |  |  |  |  |  |  |  |  |  |  |  |  |  |  |  |
| Full: 2005-2019 | 27.9 | 1.95 | 8.69 | 56.9 | 0.13 | 1.06 | 79.6 | 3.40 | 4.72 | 23.5 | 0.65 | 1.41 | 43.7 | 0.25 | 0.54 | 25.8 | NA | NA | NA |
| 1: 2005-2007 | 29.1 | 4.63 | 6.60 | 22.6 | 0.50 | 1.33 | 60.8 | 4.69 | 5.64 | 17.1 | 1.01 | 1.58 | 34.1 | 0.53 | 0.57 | 2.0 | NA | NA | NA |
| 2: 2008-2013 | 25.6 | 3.05 | 7.71 | 49.4 | 0.12 | 0.77 | 81.2 | 3.09 | 3.97 | 19.7 | 0.51 | 0.90 | 44.1 | 0.22 | 0.34 | 21.7 | NA | NA | NA |
| 3: 2014-2016 | 26.7 | 1.26 | 9.49 | 77.9 | 0.10 | 0.91 | 91.6 | 2.73 | 4.87 | 41.8 | 0.51 | 1.21 | 57.0 | 0.21 | 0.69 | 54.3 | NA | NA | NA |
| 4: 2017-2019 | 32.3 | 0.62 | 13.33 | 94.6 | 0.08 | 1.44 | 94.8 | 3.16 | 4.72 | 28.8 | 0.71 | 3.00 | 71.9 | 0.19 | 1.03 | 68.9 | NA | NA | NA |

*Acronyms: % NM = % non-missing data; OBS=observed concentrations; CF=estimated counterfactual concentrations; % Diff = 100*(CF-OBS)/CF.*

*Time periods: All=2005-2019 (n=5,478 days); 1=2005-2007 (n=1,095 days); 2=2008-2013 (n=2,192 days); 3=2014-2016 (n=1,096 days); 4=2017-2019 (n=1,095 days). Note: Most data available ~30% of days; data at NY-Bronx during Period 2 only available 9.2% of days because of site renovations (Chapter 2), and data at LA-North Main during Periods 1 and 2 only available on 15.9% and 23.1% of days, respectively, because of only 1-in-6 day sampling during 2005-2010 (Chapter 2).*

**Table S5.** Percent and daily number of respiratory visits averted during 2005-2019 in New York City, Los Angeles, and Atlanta using the primary health benefits estimation approach, by policy group and pollutant exposure set.

| **City** | **Policy Group** | **1: PM_2.5_ only** | | **2: Criteria Pollutants** | | **3: PM_2.5_ Components + Criteria Gases** | | **4: PM_2.5_ Sources** | |
| --- | --- | --- | --- | --- | --- | --- | --- | --- | --- |
|  |  | **% Averted** | **# Per Day** | **% Averted** | **# Per Day** | **% Averted** | **# Per Day** | **% Averted** | **# Per Day** |
| New York City | All | 3.0 (2.1, 3.9) | 11.2 (7.9, 14.6) | 7.1 (5.4, 8.9) | 24.7 (17.9, 31.5) | 3.4 (0.7, 6.1) | 10.1 (1.6, 18.6) | 0.8 (-0.4, 2.0) | 3.5 (-0.8, 7.9) |
|  | EGU | 2.6 (2.0, 3.2) | 7.9 (6.1, 9.7) | 5.5 (3.9, 7.1) | 16.8 (11.2, 22.5) | 5.1 (3.2, 7.0) | 14.9 (8.0, 21.8) | 0.8 (-0.1, 1.6) | 2.0 (-0.6, 4.6) |
|  | Mobile | 0.3 (0.2, 0.4) | 1.5 (1.2, 1.9) | 0.6 (0.3, 0.8) | 2.3 (1.3, 3.3) | -0.3 (-1.4, 0.8) | -0.8 (-4.2, 2.5) | 0.3 (-0.5, 1.1) | 1.6 (-1.1, 4.3) |
| Los Angeles | All | 2.1 (1.8, 2.5) | 2.6 (2.1, 3.1) | 2.4 (1.4, 3.4) | 2.7 (0.7, 4.7) | 0.6 (-0.7, 1.9) | 0.7 (-1.5, 2.9) | 2.6 (1.3, 4.0) | 2.3 (0.6, 3.9) |
|  | Mobile | 1.3 (1.1, 1.5) | 1.9 (1.5, 2.2) | 1.5 (1.2, 1.8) | 2.7 (2.1, 3.3) | 1.1 (0.5, 1.6) | 2.2 (1.3, 3.1) | 2.1 (1.5, 2.7) | 2.3 (1.6, 3.0) |
|  | Port | 0.5 (0.4, 0.6) | 0.6 (0.4, 0.7) | 0.3 (-0.5, 1.1) | -0.5 (-2.1, 1.0) | -0.4 (-1.5, 0.6) | -1.2 (-3.0, 0.7) | 1.0 (0.3, 1.7) | 1.5 (0.2, 2.7) |
| Atlanta | All | 3.5 (1.4, 5.6) | 5.0 (1.9, 8.1) | 4.5 (0.8, 8.2) | 6.4 (1.0, 11.9) | 2.1 (-1.9, 6.1) | 2.8 (-2.7, 8.4) | 1.8 (-0.9, 4.5) | 2.6 (-1.3, 6.4) |
|  | EGU | 2.6 (1.5, 3.6) | 3.6 (2.1, 5.1) | 2.3 (0.5, 4.1) | 3.2 (0.7, 5.7) | 2.9 (1.2, 4.6) | 4.0 (1.6, 6.4) | -0.2 (-1.2, 0.8) | -0.3 (-1.7, 1.0) |
|  | Mobile | 1.4 (1.0, 1.9) | 2.0 (1.4, 2.6) | 1.9 (0.7, 3.1) | 2.7 (1.0, 4.3) | 0.6 (-1.2, 2.5) | 0.8 (-1.7, 3.4) | 1.9 (0.1, 3.6) | 2.6 (0.1, 5.2) |

*Note: The primary health benefits estimation approach considered the full study period (2005-2019) as a single time period in the health benefits estimation process.*

*Note: The city-specific results are pooled estimates of the percent and number of visits averted, which represent averages for populations living within 10-mile (roughly 314 square mile) areas represented by the city-sites. The reported numbers of visits averted represent the average count for the monitor-based study populations within each city,not sums across monitors or estimates for the entire city population.*

**Table S6.** Percent and daily number of cardiovascular visits averted during 2005-2019 in New York City, Los Angeles, and Atlanta using the primary health benefits estimation approach, by policy group and exposure set.

| **City** | **Policy Group** | **1: PM_2.5_ only** | | **2: Criteria Pollutants** | | **3: PM_2.5_ Components + Criteria Gases** | | **4: PM_2.5_ Sources** | |
| --- | --- | --- | --- | --- | --- | --- | --- | --- | --- |
|  |  | **% Averted** | **# Per Day** | **% Averted** | **# Per Day** | **% Averted** | **# Per Day** | **% Averted** | **# Per Day** |
| New York City | All | 2.3 (1.6, 3.0) | 1.9 (1.3, 2.6) | 2.6 (0.9, 4.2) | 2.1 (0.7, 3.6) | 3.1 (0.5, 5.8) | 2.6 (0.3, 4.8) | 2.0 (0.7, 3.2) | 1.6 (0.5, 2.8) |
|  | EGU | 1.9 (1.4, 2.4) | 1.5 (1.1, 1.9) | 2.4 (0.9, 3.9) | 2.0 (0.7, 3.2) | 3.3 (1.3, 5.2) | 2.8 (1.1, 4.4) | 1.2 (0.4, 2.1) | 1.0 (0.3, 1.7) |
|  | Mobile | 0.3 (0.2, 0.4) | 0.3 (0.2, 0.3) | 0.7 (0.5, 1.0) | 0.6 (0.4, 0.8) | 0.4 (-0.9, 1.7) | 0.3 (-0.9, 1.4) | 0.9 (0.1, 1.8) | 0.8 (0.1, 1.5) |
| Los Angeles | All | 0.6 (0.2, 1.0) | 0.2 (0.1, 0.4) | 1.2 (0.3, 2.1) | 0.5 (0.0, 1.1) | 1.6 (0.2, 3.0) | 0.5 (-0.2, 1.2) | 1.2 (-0.2, 2.5) | 0.3 (-0.1, 0.8) |
|  | Mobile | 0.3 (0.1, 0.5) | 0.1 (0.0, 0.2) | 0.4 (0.1, 0.7) | 0.2 (0.0, 0.3) | 0.5 (-0.1, 1.1) | 0.2 (-0.1, 0.4) | 0.3 (-0.4, 1.0) | 0.1 (-0.1, 0.3) |
|  | Port | 0.1 (0.0, 0.3) | 0.1 (0.0, 0.1) | 0.4 (-0.3, 1.1) | 0.1 (-0.4, 0.5) | 0.5 (-0.6, 1.7) | 0.2 (-0.4, 0.8) | 0.5 (-0.4, 1.3) | 0.3 (-0.1, 0.6) |
| Atlanta | All | -0.1 (-2.8, 2.6) | 0.0 (-0.9, 0.8) | -0.7 (-5.5, 4.1) | -0.2 (-1.6, 1.2) | -0.1 (-6.3, 6.1) | 0.0 (-1.9, 1.9) | -1.8 (-5.3, 1.7) | -0.5 (-1.6, 0.5) |
|  | EGU | 0.5 (-0.8, 1.8) | 0.1 (-0.3, 0.6) | 0.2 (-2.1, 2.5) | 0.1 (-0.6, 0.8) | -0.7 (-3.2, 1.9) | -0.2 (-1.0, 0.6) | -0.4 (-1.7, 0.8) | -0.1 (-0.5, 0.2) |
|  | Mobile | 0.5 (0.0, 1.0) | 0.2 (0.0, 0.3) | 0.4 (-1.0, 1.9) | 0.1 (-0.3, 0.6) | 0.2 (-2.6, 3.1) | 0.1 (-0.8, 0.9) | -0.5 (-2.6, 1.7) | -0.1 (-0.8, 0.5) |

*Note: The primary health benefits estimation approach considered the full study period (2005-2019) as a single time period in the health benefits estimation process.*

*Note: The city-specific results are pooled estimates of the percent and number of visits averted, which represent averages for populations living within 10-mile (roughly 314 square mile) areas represented by the city-sites. The reported numbers of visits averted represent the average count for the monitor-based study populations within each city,not sums across monitors or estimates for the entire city population.*

Figure S1. Timeline of key air quality policies included in this study and their implementation dates.

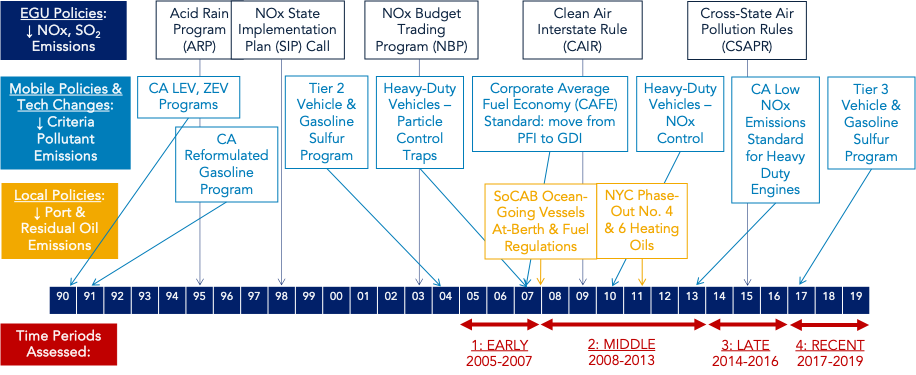
